# The Association of Long-Term Body Mass Index Variability with the Development of HFpEF and HFrEF Across Patterns of Weight Change

**DOI:** 10.1101/2024.11.08.24317010

**Authors:** Zeshui Yu, Yuqing Chen, Oshin Miranda, Meiyuzhen Qi, Manling Zhang, Ning Feng, Timothy P Ryan, Nanette Cathrin Schloot, Yu Chen, Flora Sam, Lirong Wang

## Abstract

**Background:** Recent studies have shown BMI variability is risk factor for various adverse cardiovascular outcomes. However, the specific associations between BMI variability and the risk of developing HFpEF versus HFrEF, particularly across multiple weight change trends, remain unexplored.

**Methods and Results:** We identified a cohort of 52,286 eligible patients with overweight or obesity grouped into three categories based on their BMI change patterns over five years: weight loss, stable weight, and weight gain. BMI variability was assessed in the same 5-year period using average successive variability (ASV). These patients were subsequently followed to monitor the incidence of HFpEF and HFrEF. Cox regression models were applied to examine the differential association between BMI variability and HFpEF and HFrEF risk. Over a median follow-up of 4.81 years, 2,295 patients developed HFpEF, and 1,189 developed HFrEF. After adjusting for relevant confounders, elevated BMI variability was associated with an increased risk of HFpEF. The hazard ratios (HRs) of HFpEF for each 1-SD increment in ASV of BMI were 1.27 (95% CI, 1.10-1.47) in the weight loss group and 1.22 (95% CI, 1.09-1.37) in the stable weight group. Additionally, when analyzed as a binary variable divided by the median, BMI variability above the median was associated with higher risks of HFpEF compared to those below the median, with the corresponding HRs being 1.46 (95% CI, 1.20-1.77) for the weight loss group and 1.17 (95% CI, 1.04-1.31) for the stable weight group.

**Conclusions:** In this large cohort of patients living with overweight or obesity, greater BMI variability was significantly associated with a higher risk of developing HFpEF compared to patients with reduced and stable weight over time.

**Clinical Perspective What’s new?:**

1. In patients with weight loss and stable weight, those with higher BMI variability have an increased risk of developing incident HFrEF compared to those experiencing lower BMI variability, after adjusting all potential confounding variables.
2. In patients with weight gain, BMI variability was not significantly linked to the risk of developing HFpEF or HFrEF. However, a larger increase in delta BMI was significantly associated with a higher risk of incident HFpEF and HFrEF in this group.

**What are the clinical implications?:**

1. Promoting the importance of stable and consistent weight management strategies to reduce heart failure risk, particularly by minimizing BMI variability in patients undergoing weight loss or maintaining stable weight.

## Introduction

Heart Failure (HF) is a major public health problem, with an estimated global prevalence of >64 million adults^1–3^. Similarly, the prevalence and incidence of HF continues to increase in the United States, affecting around 6.2 million adults. In the HF population, HF with preserved ejection fraction (HFpEF) accounts for > 50%, compared to HF with reduced ejection fraction (HFrEF), with the proportion of HFpEF relative to HFrEF increasing at a concerning rate of 1% per year.^4–8^

Obesity-associated cardiometabolic disorders in an aging population are accelerating and are associated with increased cardiovascular morbidity and mortality. The obesity rate is expected to be around 50% by 2035 in aging populations but this is likely an underestimation, given that the burgeoning prevalence of overweight and obesity among children and adolescents is not considered in this calculation.^9^ Indeed, almost 40 million children under 5 years and 340 million between 5 and 19 years old are estimated to be obese or overweight worldwide.^10^

There remains considerable heterogeneity in the trajectories of weight change among individuals with overweight and obesity.^11,12^ Distinct patterns of weight change differentially impact cardiovascular health.^13–16^ Weight loss often reflects either exacerbation of existing illness or adherence to the lifestyle intervention, while weight gain frequently relates to worse metabolic profiles or physiological responses to the pharmacological treatment. To gain a comprehensive understanding of the pathophysiology of heart failure in populations with overweight or obesity, it is crucial to group patients according to their weight change patterns: those who lose weight, those who maintain a stable weight, and those who gain weight. Prior studies have demonstrated that both weight loss and weight gain independently increase the risk of various cardiovascular outcomes, highlighting the importance of distinguishing between these different weight change patterns when investigating their impact on heart failure subtypes.

Despite emerging evidence associating visit-to-visit Body Mass Index (BMI) variability with various cardiovascular outcomes, including overall heart failure, the limited integration of BMI variability into clinical risk assessment arises from a lack of compelling evidence from real-world clinical data.^17–19^ Additionally, limited information is known about the differential association between BMI variability and the outcomes of incident HFpEF versus HFrEF in patients with overweight or obesity. Similarly, HFpEF and HFrEF likely have distinct pathophysiological mechanisms that underlie each respective obesity phenotype. Evidence suggests that obesity may be a stronger risk factor for HFpEF, particularly in women. ^20^ We sought to investigate the association between long-term BMI variability and the risk of developing HFpEF versus HFrEF in patients who are overweight and obese, without prevalent HF at baseline, using longitudinal clinical data collected from an academic hospital setting in Pennsylvania.

## METHODS

We randomly selected 100,000 patients with overweight and obesity who had an initial BMI record equal to or greater than 27 kg/m^2^ on an index date and were followed for at least 5 years between January 1^st^,2004, and October 31^st^, 2021, in the University of Pittsburgh Medical Center (UPMC). Of these individuals, the 100,000 patients were divided into four equal groups of 25,000 subjects based on their initial BMI values. The groups were: Overweight (27 kg/m^2^<=BMI<30 kg/m^2^), class I obesity (30 kg/m^2^<=BMI<35 kg/m^2^), class II obesity (35 kg/m^2^<=BMI<40 kg/m^2^) and class III obesity (BMI>=40 kg/m^2^). This BMI record on the index date is defined as the **initial BMI** value, which represents the earliest BMI measurement that meets the criteria of the individual patient’s group and serves as the beginning of the BMI variability measurement phase. The BMI variability measurement period was defined as the five years following the initial BMI value, using BMI records from EHRs during this 5-year period to create an independent variable representing each patient’s BMI variability. As shown in **Figure 1**, the **baseline** was defined as the end of the BMI variability measurement period (i.e., 5 years), from which incident HF was then evaluated. 2,807 patients were excluded from this analysis because of data input errors. Patients were excluded if they did not have at least one BMI record per calendar year during the 5-year BMI variability measurement period (n=18,468). Given our focus on weight change patterns and BMI variability, this criterion was applied to ensure consistent and continuous BMI measurement throughout the study. Other exclusion criteria included a prior history of bariatric surgery (n=2,647) and cancer (n=11,654), prevalent HF during the BMI variability measurement period of 5 years (i.e. Before the baseline (n=8,793)), missing covariates (n=2,446), and subsequent unclassified HF during the follow-up period (n=953) (Figure 2)

**Figure 1:**
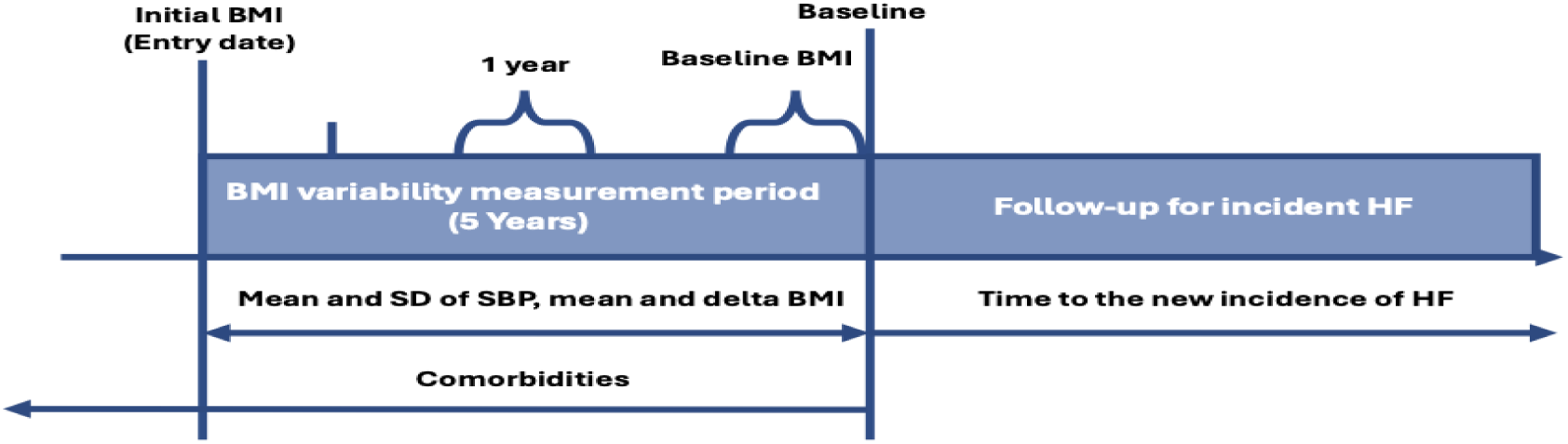
Study design scheme. The initial BMI value represents the earliest available BMI record that fall into the range of each group and serves as the index (i.e., the starting point of the BMI variability measurement period). The baseline of the study was established at the end of this BMI variability measurement period (i.e., the ending point of the BMI variability measurement period and the beginning of the follow-up period). From this point forward, the occurrence of heart failure was monitored until the earliest occurrence of a HF diagnosis or HF event, death, the last encounter date, or October 31st, 2021, whichever occurred first. Abbreviation: BMI=body mass index; HF=heart failure; SD=standard deviation; SBP=systolic blood pressure.

**Figure 2:**
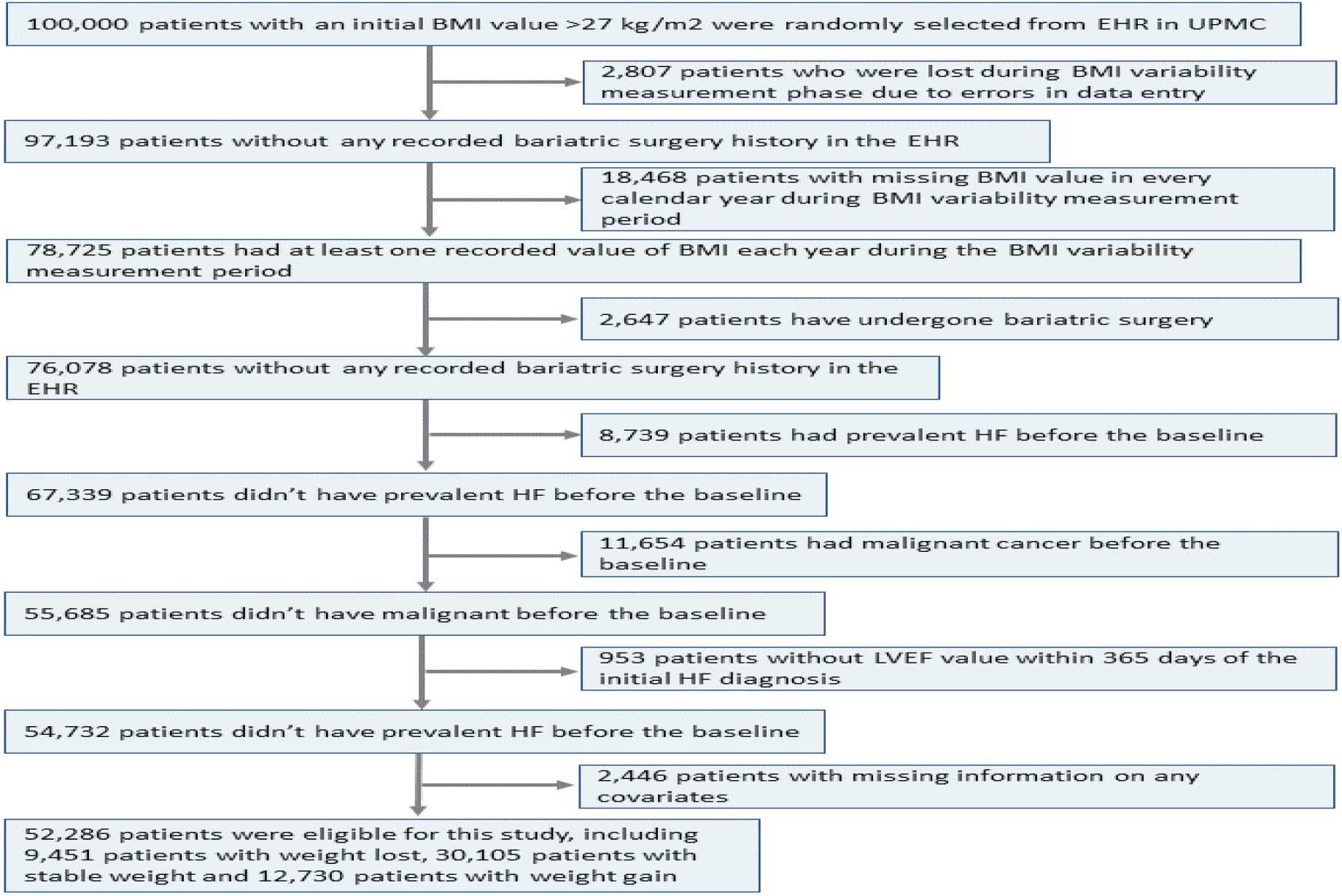
Flow Diagram for Included Patients. This flowchart summarizes the clinical and data inclusion/exclusion criteria applied in the study. Abbreviation: BMI=body mass index; EHR=electronical health record; HF=heart failure; ASCVD=atherosclerotic cardiovascular disease; AFIB=atrial fibrillation; T2DM=type 2 diabetes mellitus; LVEF=left ventricle ejection fraction.

### Measurement of variability in BMI

We calculated individual-level BMI variability during the measurement period using Average Successive Variability (ASV) as described in the study by Petria *et al*.^21^ ASV is the mean of absolute differences between consecutive BMI measurements. This method emphasizes time-sensitive changes by focusing on sequential changes as well as overall variability. ASV was selected as the variable of interest because a prior systematic review and meta-analysis revealed that it is the most frequently employed measure for assessing BMI variability in the current body of published research.^16,19^

### Definition of incident heart failure subtypes

The incidence of new-onset HF was identified as the first-time diagnosis of clinical HF as classified by ICD-9 and ICD-10 codes during the follow-up. ICD-9 and ICD-10 are code sets for classifying diseases and health problems (Table S1). Patients were followed from baseline to the earliest occurrence of a HF diagnosis or HF event, death, the last encounter date, or October 31st, 2021, whichever occurred first. The measure of left ventricle ejection fraction (LVEF) was obtained from the most recent echocardiogram test within a year of incident HF and was used to determine HF subtypes. HFpEF was defined as LVEF > 50%, while HFrEF was defined as LVEF ≤ 50%.^22^

### Covariates

Covariates included gender, race, baseline age, smoking status, the number of BMI records and mean BMI during the measurement period, baseline BMI categories, and comorbidities (atherosclerotic cardiovascular disease, atrial fibrillation, type 2 diabetes mellitus, hypertension, hyperlipidemia, chronic kidney disease, nonalcoholic fatty liver disease/nonalcoholic steatohepatitis, and obstructive sleep apnea). The mean and standard deviation of heart rate and systolic blood pressure (SBP) during the BMI variability measurement period were included. Additionally, delta BMI changes were measured by the difference between the initial BMI and the baseline BMI over the BMI variability measurement period.

### Statistical analysis

Patient characteristics were summarized as means ± standard deviations for continuous variables and counts (percentages) for categorical variables. Baseline demographics and characteristics were compared across three groups of participants presenting with distinct patterns of BMI change: weight loss (delta BMI change of < −5%); stable weight (delta BMI change between ≥-5 and +5%) and weight gain, (delta BMI change of > +5%). The threshold of 5% weight loss is adopted based on evidence indicating improved cardiometabolic traits and cardiovascular outcomes in patients achieving it and is commonly used as the definition of clinically significant weight loss in clinical trials.^23–26^ Statistical comparisons were performed using the Chi-square test for categorical variables and ANOVA for continuous variables among these groups. The granular percentage weight change patterns were analyzed alongside the percentage of patients within each initial BMI category. The percentage of weight change was calculated and classified into the following categories: −10% or less, >-10% to −5%, >-5% to - 2.5%, >-2.5% to 0%, >0% to 2.5%, >2.5% to 5%, >5% to 10%, and 10% or more.

We first conducted a Cox regression analysis on the entire study cohort to explore the relationship between 5-year BMI variability and the risk of developing either HFpEF or HFrEF during the follow-up phase. Following this, we further analyzed three distinct groups categorized by BMI change patterns, acknowledging the significant heterogeneity among patients with overweight and obesity. The Cox regression models accounted for the presence of other types of HF and death as competing risks, providing adjusted hazard rates for the occurrence of one type of HF influencing the censoring of the other type. BMI variability, measured by ASV, were examined as continuous values, as well as medians and quartiles. Hazard ratios (HRs) were reported for each standard deviation increase in continuous measures and compared across categories, using the below-median group or the lowest quartile as the reference. All models were adjusted for the covariates listed above. Incidence rates of heart failure were determined by dividing the total number of patients diagnosed with first-time heart failure by the total person-years accumulated during the follow-up period. We also sought to conduct analyses to assess the gender differences in the relationship between BMI variability and the incidence of HFpEF and HFrEF across different weight change patterns.

Furthermore, we assessed the association between baseline BMI categories and the incidence of HF subtypes according to different BMI change patterns. These analyses were conducted using cause-specific Cox proportional hazard models in three separated BMI delta-change pattern groups, adjusted for age, gender, race, smoking status, the number of BMI records, and all comorbidities. Statistical significance was established at a two-sided p-value of less than 0.05. All statistical analyses were performed using Python 3.9 and SAS software.

## RESULTS

During the mean follow-up period of 4.81 years, a total of 3,484 patients had a new diagnosis of HF, with 2,295 cases classified as HFpEF and 1,189 cases classified as HFrEF. Table 1 displays the baseline demographics and clinical characteristics of the study patients by BMI change pattern. In comparison with patients with weight loss or stable weight, patients with weight gain were younger (53.19 years old, SD: 15.13). Female patients were more likely to experience either weight gain or weight loss compared to male patients. Those who experienced weight loss have higher rates of ASCVD, AFIB, CKD, HTN, HLP, NAFLD, and T2DM at the baseline compared to patients with stable weight or those who gained weight. Conversely, patients who gained weight have the highest rate of OSA. Patients with weight gain and weight loss exhibit higher average SD of heart rate, blood pressure, and BMI variability compared to those with stable weight. Differences in baseline demographics and characteristics of the study cohort based on quartiles of ASV and HF subtypes are provided in Tables S2 and S3 in the Supplement.

**Table 1.**
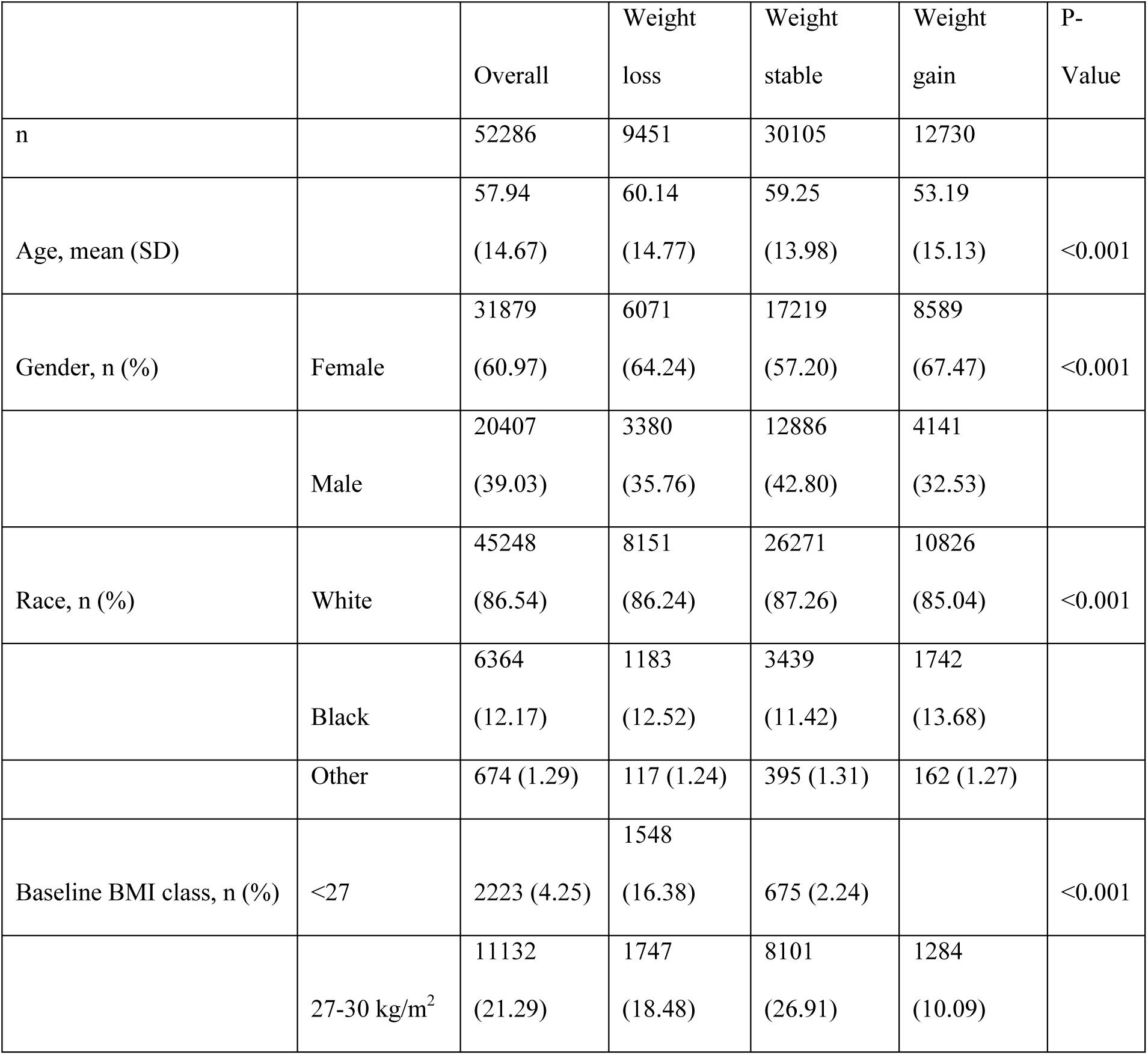
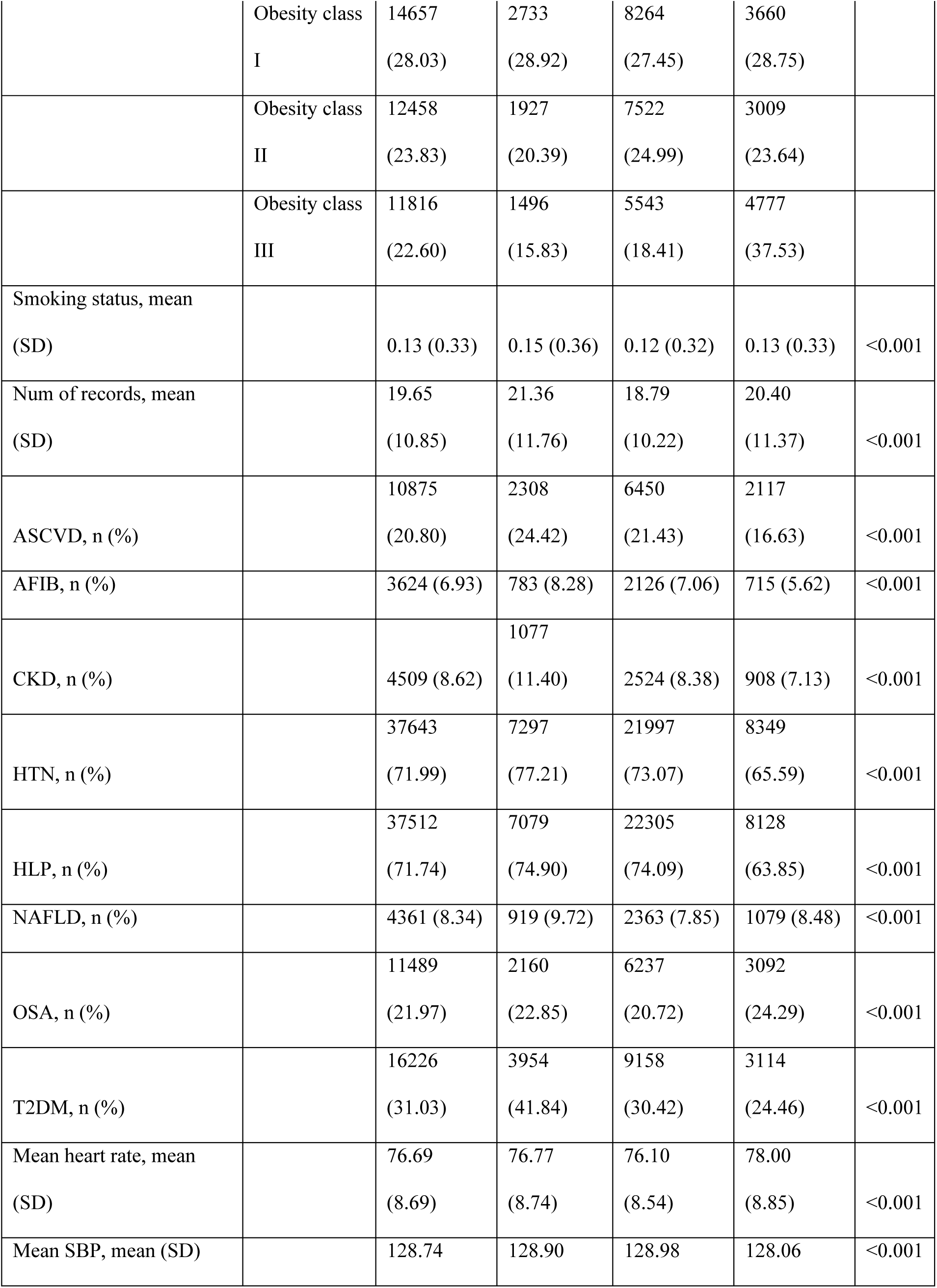
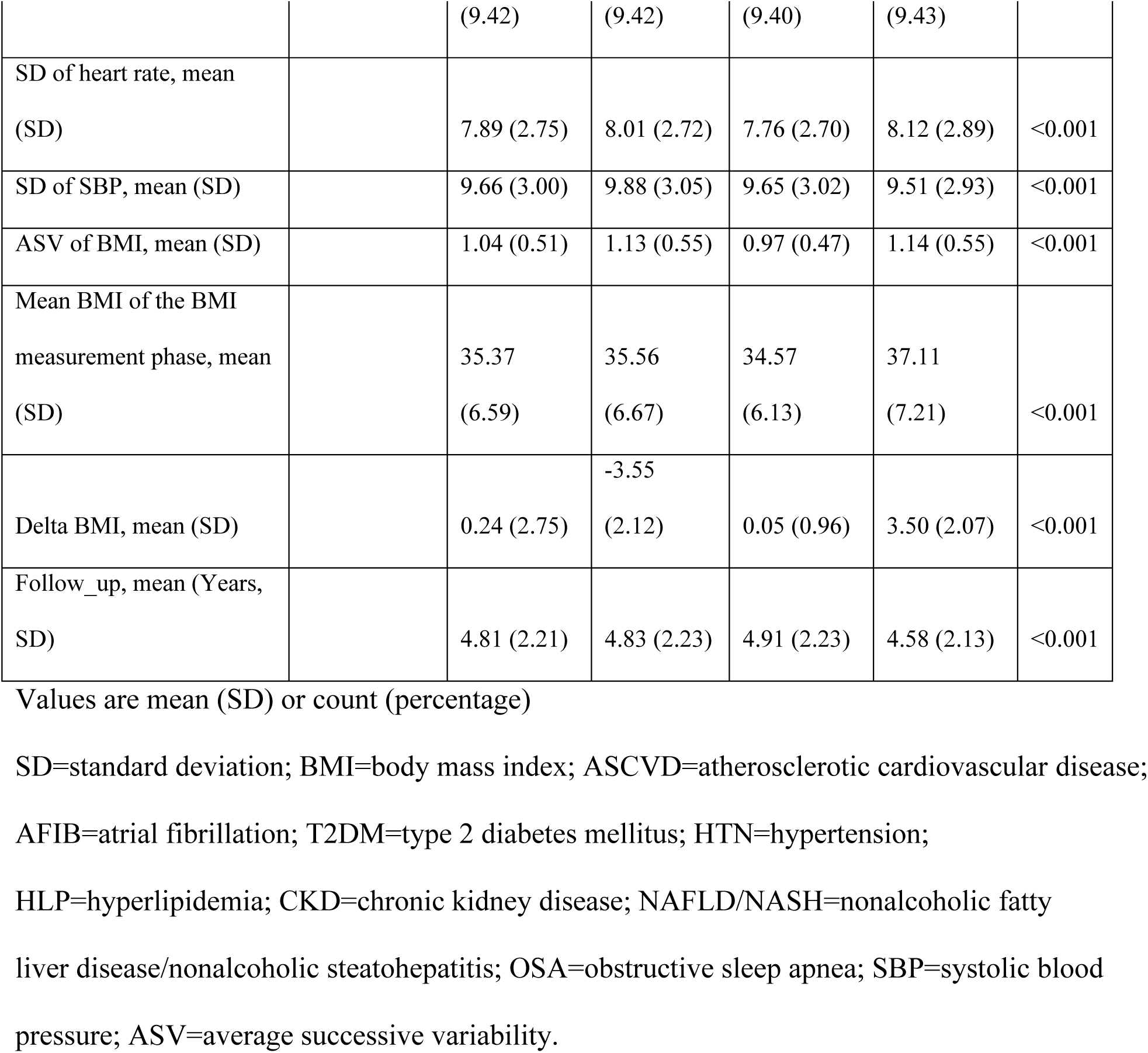
Demographic and Baseline Characteristics of Patients Categorized by BMI Change Patterns.

The granular patterns of weight change were examined in relation to the corresponding percentage of patients within each initial BMI categories. Figure 3 illustrates the proportion of patients within these groups. By the end of the 5-year measurement phase, patients in the Obesity Group III category at the index date were more likely to experience a weight loss or gain of more than 10%. Conversely, a higher proportion of patients in the overweight category maintained stable weight over the same period.

**Figure 3.**
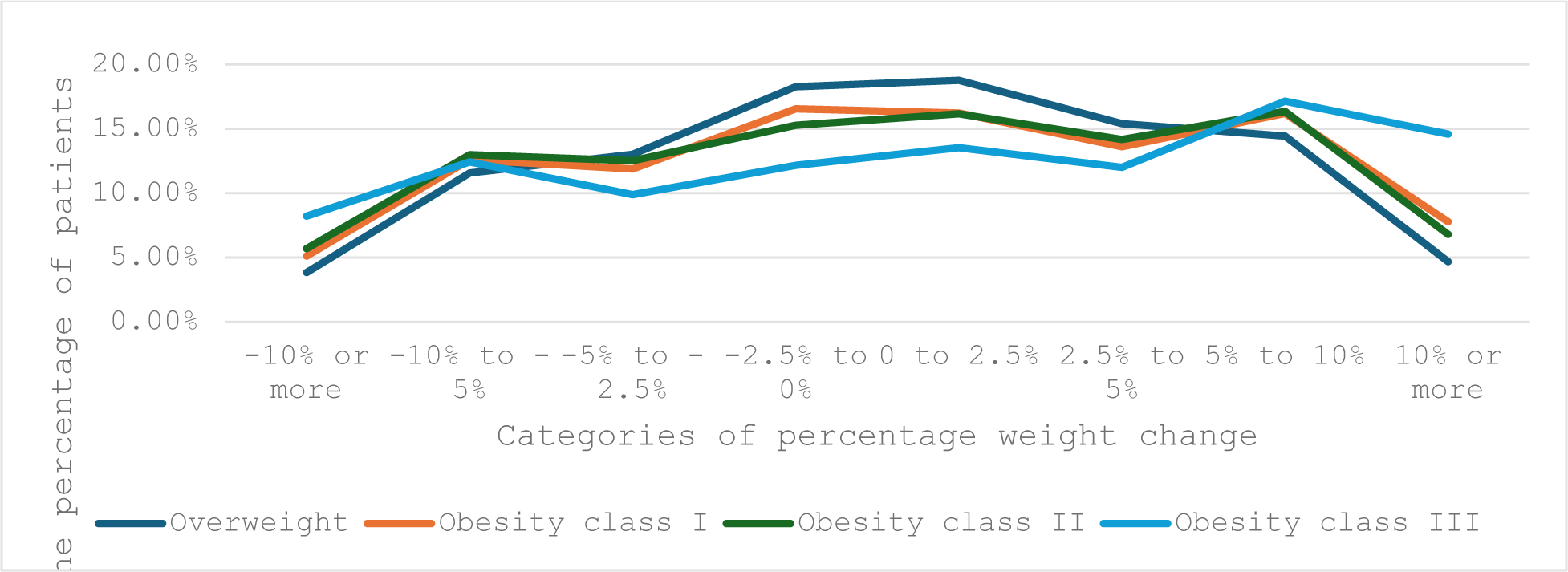
Percentage of Patients with Different Weight Changes Across Initial BMI Categories.

### THE ASSOCIATION BETWEEN BMI VARIABILITY AND HF SUBTYPES

Table 2 presents the adjusted HRs of ASV of BMI for the incident HFpEF and HFrEF in the entire study cohort and within groups of three patterns of BMI changes. In the overall study cohort, a significant association was found between increasing BMI variability and the risk of incident HFpEF and HFrEF. The adjusted HRs for incident HFpEF were 1.26 (95% CI: 1.17-1.36), 1.21 (95% CI: 1.11-1.33), and 1.37 (95% CI: 1.21-1.56), and for incident HFrEF were 1.13 (95% CI: 1.00-1.27), 1.18 (95% CI: 1.04-1.33), and 1.30 (95% CI: 1.09-1.54), corresponding to BMI variability (ASV) as a continuous variable, and the higher median and highest quartile compared to the lower median and lowest quartile, respectively, in the overall study cohort.

**Table 2.**
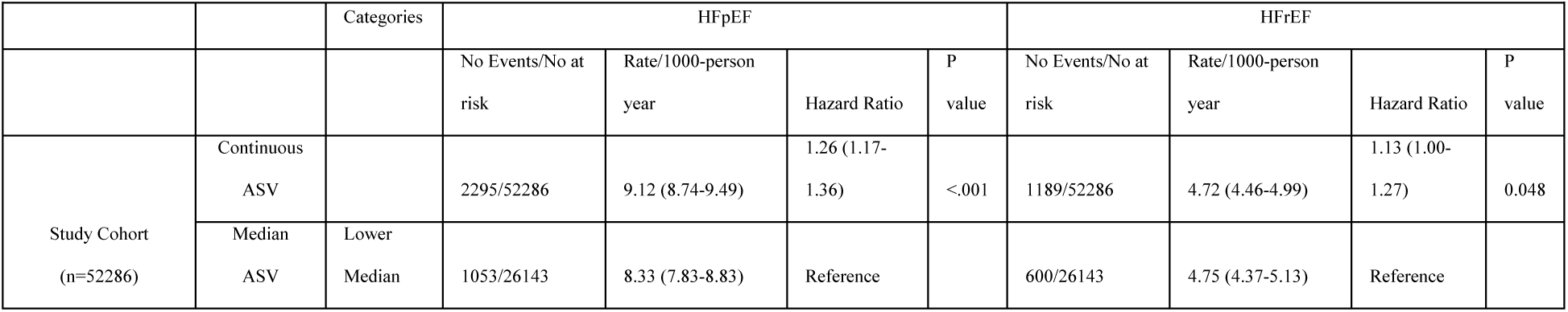
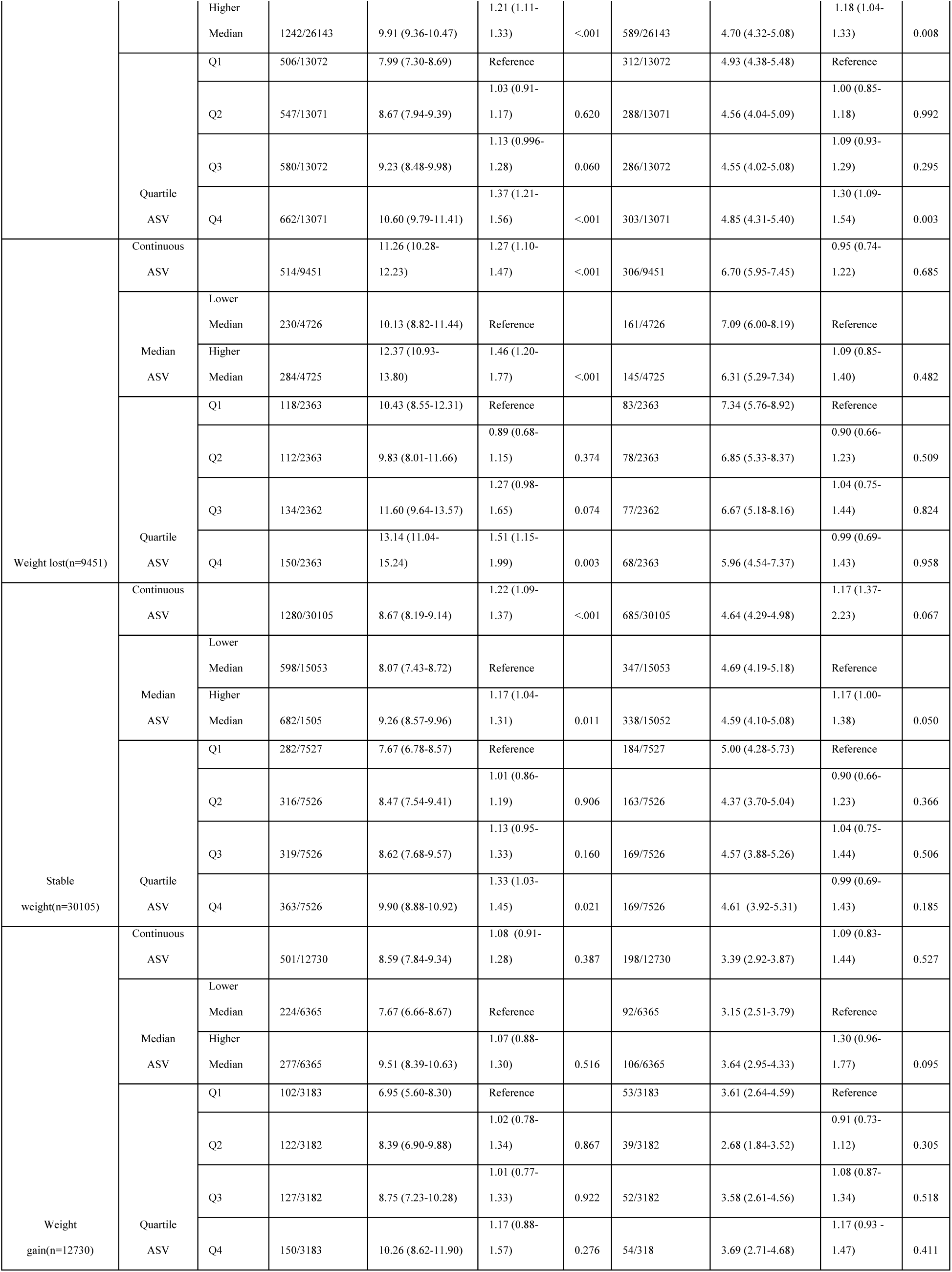
Association of BMI ASV with Incident HFpEF and HFrEF in the Study Cohort and Across Weight Change Patterns: The models were adjusted for age, gender, race, smoking status, and comorbidities (atherosclerotic cardiovascular disease, atrial fibrillation, hypertension, hyperlipidemia, chronic kidney disease, nonalcoholic fatty liver disease/nonalcoholic steatohepatitis, obstructive sleep apnea, and type 2 diabetes mellitus), the mean and standard deviation of heart rate and systolic blood pressure and delta weight change. BMI=body mass index; HFpEF=heart failure with preserved ejection fraction; HFrEF=heart failure with reduced ejection fraction.

When the study cohort was grouped by weight change patterns, continuous BMI variability was positively associated with an increased risk of developing HFpEF in both the weight loss and weight gain groups. The HRs of HFpEF for each 1-SD increment in ASV of BMI were 1.27 (95% CI, 1.10-1.47) in the weight loss group and 1.22 (95% CI, 1.09-1.37) in the stable weight group. However, in the group of weight gain, there was no significant association between BMI variability and incident HFpEF or HFrEF.

When BMI variability was measured as a categorical variable, patients in the above-median BMI variability group had an increased risk of incident HFpEF compared to those below the median, with HRs of 1.46 (95% CI, 1.20-1.77) for the weight loss group and 1.17 (95% CI, 1.04-1.31) for the stable weight group. Additionally, in the weight loss group patients in the fourth quartile had a higher risk of incident HFpEF compared to those in the first quartile, with HRs of 1.51 (95% CI, 1.15-1.99) for the stable weight group it was 1.33 (95% CI, 1.03-1.45). No significant associations were observed between categorical BMI variability and incident HF in the weight gain group. However, a greater delta BMI increase within this group was significantly associated with a higher risk of incident HFpEF and HFrEF, with HRs of 1.09 (95% CI, 1.04– 1.13) and 1.09 (95% CI, 1.00–1.18) for HFpEF and HFrEF, respectively, when BMI variability was measured as a continuous variable.

Importantly, there was no significant association between ASV of BMI and incident HFrEF across any weight change pattern of BMI. In the gender-specific effect analyses, results for both female and male patients showed no significant differences, with no notable interaction between gender * continuous BMI variability in the weight loss group (Table S4).

### THE ASSOCIATION OF BASELINE BMI WITH HFpEF and HFrEF

We examined the relationship between baseline BMI categories and the risk of HF subtypes as reported in Table 3. Patients with severe obesity at baseline, defined as Obesity Class III, exhibited a significantly increased risk of incident HFpEF across all three subgroups. Specifically, the HRs for Obesity Class III, compared to those with a BMI less than 27, were 2.21 (95% CI, 1.59-3.07) in the weight loss group and 2.13 (95% CI, 1.42-3.17) in the stable weight group. Furthermore, the HR for HFpEF associated with Obesity Class III relative to Obesity Class I was 4.47 (95% CI, 2.84-7.03). Patients in Obesity Class II displayed a 1.58-fold increased risk of incident HFpEF compared to those with a BMI less than 27 in the weight loss group. Those in Obesity Classes I and II faced higher risks for incident HFpEF compared to patients categorized as overweight in the weight gain group, with HRs of 2.07 (95% CI, 1.30-3.29) and 2.49 (95% CI, 1.57-3.96), indicating a trend of escalating HRs with higher obesity classes.

**Table 3.** The Association of Baseline BMI Categories with the Risk of Incident HFpEF vs HFrEF by Weight Change Patterns: The models were adjusted for age, gender, race, smoking status, and comorbidities (atherosclerotic cardiovascular disease, atrial fibrillation, hypertension, hyperlipidemia, chronic kidney disease, nonalcoholic fatty liver disease/nonalcoholic steatohepatitis, obstructive sleep apnea, and type 2 diabetes mellitus) BMI=body mass index; HFpEF=heart failure with preserved ejection fraction; HFrEF=heart failure with reduced ejection fraction.

| **Table 3. The Association of Baseline BMI Categories with the Risk of Incident HFpEF vs HFrEF by Weight Change Patterns** |  |  |  |  |  |
| --- | --- | --- | --- | --- | --- |
|  | BMI Categories | HFpEF |  | HFrEF |  |
|  |  | Hazard Ratio | P value | Hazard Ratio | P value |
| Weight lost(n=9451) | $\leq 27 \text{ kg/m}^2$ | reference | reference | reference | reference |
| | 27-30 $\text{kg/m}^2$ | 0.83 (0.60-1.16) | 0.273 | 1.21 (0.83-1.77) | 0.319 |
|  | Obesity class I | 1.23 (0.93-1.64) | 0.154 | 0.92 (0.63-1.34) | 0.615 |
|  | Obesity class II | 1.58 (1.17-2.14) | 0.003 | 1.49 (1.01-2.20) | 0.047 |
|  | Obesity class III | 2.21 (1.59-3.07) | <0.001 | 1.54 (0.98-2.42) | 0.06 |
| Stable weight(n=30105) | $\leq 27 \text{ kg/m}^2$ | reference | reference | reference | reference |
| | 27-30 $\text{kg/m}^2$ | 0.86 (0.58-1.28) | 0.458 | 0.92 (0.57-1.49) | 0.734 |
|  | Obesity class I | 1.08 (0.73-1.60) | 0.696 | 0.99 (0.61-1.61) | 0.981 |
|  | Obesity class II | 1.38 (0.93-2.05) | 0.111 | 1.10 (0.67-1.79) | 0.717 |
|  | Obesity class III | 2.13 (1.42-3.17) | <0.001 | 1.75 (1.06-2.90) | 0.029 |
| Weight gain(n=12730) | 27-30 kg/m<sup>2</sup> | reference | reference | reference | reference |
|  | Obesity class I | 2.07 (1.30-3.29) | 0.002 | 0.98 (0.61-1.59) | 0.946 |
|  | Obesity class II | 2.49 (1.57-3.96) | <0.001 | 1.00 (0.60-1.65) | 0.989 |
|  | Obesity class III | 4.47 (2.84-7.03) | <0.001 | 1.21 (0.74-2.00) | 0.444 |

Additionally, in the weight loss group, patients with Obesity Class II were associated with an increased risk of HFrEF compared to those with a BMI less than 27, with an HR of 1.49 (95% CI, 1.01-2.20). In the stable weight group, patients in Obesity Class III had a higher risk for HFrEF compared to those with a BMI less than 27, with an HR of 1.75 (95% CI, 1.06-2.90). No significant association between baseline BMI and HFrEF was observed in the weight gain group.

## DISCUSSION

A comprehensive study was conducted to examine the relationship between long-term BMI variability and the risk of developing HFpEF and HFrEF in a large cohort of overweight and obese individuals, with particular consideration given to their weight change patterns. The primary findings revealed that greater BMI variability was independently associated with an increased risk of both HFpEF and HFrEF within this cohort. These associations remained significant even after adjusting for all potential confounding factors, including baseline BMI and changes in BMI over time. Furthermore, the analysis demonstrated that the link between BMI variability and future HFpEF risk persisted in patients with specific weight loss or stable weight patterns. Among patients with weight gain, BMI variability was not significantly associated with the risk of incident HFpEF or HFrEF; however, a greater magnitude of delta BMI increase was significantly linked to an increased risk of incident HFpEF and HFrEF in this group. These findings expand the previous evidence that has focused primarily on the association between BMI variability and cardiovascular disease and overall HF.^17–19^ Our findings contribute to a deeper understanding of the complex mechanisms of HF subtypes and may inform obesity management strategies.

To our knowledge, our study is the first study to explore the link between BMI variability with incident HFpEF and HFrEF. Several previous studies provided evidence of the association of weight variability on the risk of overall HF, mostly in patients with T2DM, although granular analyses focusing on HF subtypes were not performed due to the absence of non-invasive imaging data, such as echocardiogram or cardiac MRI. ^17,18,27,28^ The availability of echocardiogram report in this study permitted accurate identification based on the specific subtype of incident HF cases. It is important to note that HF is a clinical syndrome and cannot be solely identified by a normal LVEF. Importantly, in our study, patients were selected from hospital EHRs and identified cases with a confirmed diagnostic assessment of HF based on ICD coding. Therefore, we addressed the current gap in evidence regarding the relationship between BMI variability and the risks of HFpEF and HFrEF in a large cohort of individuals with obesity and overweight over an extended period. Additionally, BMI variability was captured using frequent measurements, averaging 19.69 records per person, collected across various clinical settings (inpatient and outpatient visits). This approach provided a detailed and accurate picture of weight changes and BMI variability over time. Our finding that BMI variability significantly increases the risk of developing HFpEF and HFrEF is in line with previous studies that have shown a positive relationship between BMI variability and cardiovascular diseases, including HF in general.^17–19^

The findings of this study have significant clinical implications for both healthcare providers and overweight or obese individuals. In clinical settings, weight reduction counseling is commonly recommended to control risk factors for obesity-related diseases and mortality.^27^ Previous studies have reported that BMI variability, regardless of the direction of weight change, can predict adverse cardiovascular events.^17,18^ However, obesity and weight change patterns differ among individuals due to various factors like genetics, unsustainable diets, psychological influences, metabolic rate changes, medication effects, medical procedures, or hormonal changes. Our approach enables a more granular analysis of how distinct weight change patterns—such as weight loss, stable weight, and weight gain—impact the risk and progression of HFpEF and HFrEF in patients with varying BMI trajectories. By addressing each group’s unique weight trajectory, this approach offers valuable insights into patient outcomes and informs the development of tailored strategies for managing cardiovascular risks associated with BMI variability. Our findings from this design offer real-world evidence underscoring the importance of minimizing body weight fluctuations when attempting to lose weight or maintain a stable weight, thereby mitigating potential adverse cardiovascular outcomes.

Moreover, as GLP-1 receptor agonists are increasingly used alongside lifestyle changes for obesity management, our findings provide valuable supportive evidence for consistent adherence to these treatments for stable and continuous weight loss. Our study suggests that reducing weight cycling could help achieve the optimal effectiveness of intervention in preventing HF.

The patient characteristics of our randomly selected group of patients align with the widely recognized pattern of higher obesity rates among women. Our study also confirmed that baseline BMI is a risk factor for HF, with a strong correlation to HFpEF than to HFrEF.^20^ This association with HFpEF is particularly pronounced in patients who gain weight gain, indicating that a higher BMI increases the risk of developing HFpEF. Interestingly, there was no significant association between BMI variability and the onset of either HFpEF or HFrEF in the weight gain group.

The exact mechanisms between BMI variability and HF, particularly HFpEF, remain incompletely understood. These mechanisms are likely complex and involve multiple factors, especially considering the prevalence of comorbidities and the markedly different outcomes of HFpEF and HFrEF to standard medical treatments. One potential mechanism may involve the long-term effects of body weight fluctuation on the development and progression of metabolic syndrome, as patients with metabolic syndrome are known to have a substantially higher risk of developing HFpEF.^29–32^ Another potential pathway could involve chronic, low-grade systemic inflammation, as evidenced by higher concentrations of C-reactive protein in patients who experience large weight fluctuations.^33^ Indeed, preclinical research has demonstrated that weight cycling (i.e. weight loss followed by weight gain) is associated with impaired energy hemostasis and a rise in hunger, and fall in satiety hormones, resulting in adipocyte hyperplasia and maladaptive excess visceral fat accumulation.^34^ Thus, the adipocyte hyperplasia and weight regain associated with weight cycling enhance the inflammatory capacity of the adipose tissue mass. Further research needs uncover precise mechanisms.

The present study has the following limitations. 1.) It is unknown whether the observed weight changes in patients were intentional or unintentional. This distinction is important because intentional weight loss from lifestyle changes is seen beneficial, while unintentional weight loss often indicates serious illness or metabolic disorder 2.) This study specifically focused on classifying the HF by ejection fraction but other HF classifications, such as New York Heart Failure classification, acute vs chronic, ischemic vs non-ischemic, or primary myocardial disease vs secondary neurohormonal activity were not available, due to the lack of data.^35^ 3.) As this was a retrospective observational study, causal inferences cannot be drawn from the results. Additionally, no significant association was found between BMI variability and the incidence of HFrEF across any weight change pattern, which may be attributed to the high heterogeneity among patients with overweight or obesity and the HFrEF phenotype. 4.) EHRs often contain entry errors, leading to the underdiagnosis of certain comorbidities.

## CONCLUSION

In this retrospective study of a large cohort of overweight and obese patients, we observed that greater variability in BMI was associated with a higher risk of developing HFpEF and HFrEF, particularly with HFpEF in patients who maintained a stable weight or lost weight. Our findings suggest that these patients should minimize weight cycling while attempting to reduce or maintain their body weight.

## Data Availability

The data used in this study were obtained from electronic health records (EHR), which contain confidential patient information. Due to privacy regulations and institutional policies, these data are not publicly available. Access to the EHR data is restricted and subject to approval by the relevant ethics and compliance committees. Researchers interested in accessing similar data are encouraged to contact the institution for information on data-sharing policies and procedures.

## Acknowledgment

ZY played a leading role in defining the research question, designing the study, analyzing the data, applying statistical methods and composing the manuscript, while also providing insights throughout the research process. YQ and MQ were involved in the study design, statistical method, and manuscript reviewing. OM contributed to manuscript reviewing. LW and YC were involved in the concept, data acquisition, study design, interpretation, comprehensive review and revision of the final manuscript, the final approval of the manuscripts, and project management. MZ and FN engaged in concept, study design, and discussion. TPR, NCS, and FS contributed to the study design, interpretation, discussion, review, and revision of the manuscript and final approval.

## Notes

### Competing Interest Statement

It is important to acknowledge the potential conflict of interest among the contributors to this research. ZY, MZ, NF, and LW report receiving research funding from Eli Lilly and Company within the past 12 months. TPR, NCS, YC, and FS are employees of Eli Lilly and Company and own company stock while the research was being conducted.

### Funding Statement

This study was funded by Eli Lilly and Company.

### Author Declarations

The University of Pittsburgh institutional review board approved this study, and informed patient consent was waived.

